# Lessons learnt from 288 COVID-19 international cases: importations over time, effect of interventions, underdetection of imported cases

**DOI:** 10.1101/2020.02.24.20027326

**Authors:** Francesco Pinotti, Laura Di Domenico, Ernesto Ortega, Marco Mancastroppa, Giulia Pullano, Eugenio Valdano, Pierre-Yves Boëlle, Chiara Poletto, Vittoria Colizza

## Abstract

288 cases have been confirmed out of China from January 3 to February 13, 2020. We collected and synthesized all available information on these cases from official sources and media. We analyzed importations that were successfully isolated and those leading to onward transmission. We modeled their number over time, in relation to the origin of travel (Hubei province, other Chinese provinces, other countries) and interventions. We characterized importations timeline to assess the rapidity of isolation, and epidemiologically linked clusters to estimate the rate of detection. We found a rapid exponential growth of importations from Hubei, combined with a slower growth from the other areas. We predicted a rebound of importations from South East Asia in the upcoming weeks. Time from travel to detection has considerably decreased since the first importation, however 6 cases out of 10 were estimated to go undetected. Countries outside China should be prepared for the possible emergence of several undetected clusters of chains of local transmissions.

## INTRODUCTION

Twenty-six countries worldwide have declared cases of the novel coronavirus, COVID-19, as of February 20, 2020^1^. Only China so far registered a widespread epidemic^2^, and authorities have implemented massive intervention measures to curtail it^3^. Outside China, affected countries are facing importations of cases and clusters of local transmission^1,4,5^ Border controls have been reinforced in many countries, and active surveillance has been intensified to rapidly detect and isolate importations, trace contacts and isolate suspect cases^6,7^.

The effectiveness of such measures, however, critically depends on COVID-19 epidemiology and natural history^8,9^, as well as the volume of importations^6^. The presence of an incubation period, during which infected individuals carry on their usual activities (including travel), is a major challenge for screening controls at airports^8^. Moreover, mild non-specific symptoms and transmission before the onset of clinical symptoms^2,10^ may compromise infection control measures for importations and onward transmissions^9^. There is concern that imported cases may have gone undetected and contribute unknowingly to the global spread of the disease^11–15^.

Here we systematically collected and analyzed data on 288 COVID-19 confirmed cases outside China. We analyzed importations that were successfully isolated and those leading to onward transmission, characterizing their case timeline. We developed a statistical model to nowcast trends in importations and quantify the proportion of undetected imported cases.

## METHODS

### Data collection and synthesis

We collected all international cases confirmed by official public health sources in the period from January 3 to February 13, 2020. Case history was reconstructed by searching the scientific literature, official public health sources, and news. Case history included: dates of travel and symptoms onset, date of COVID-19 confirmation, date of hospital admission, date of case isolation, travel history, epidemiological link with other cases, hospitalization history. International cases included imported cases, secondary cases out of China, and repatriations. Cases from cruises were not considered here. Information was extracted by LDD and EO and checked by MM. The full database, along with the database describing clusters, were made publicly available^16^.

### Descriptive analysis

For imported cases with full information on the timeline of events, we computed the average duration from travel to onset, from travel to hospitalization, and from hospitalization to reporting. We used analysis of variance to compare groups of imported cases that generated or did not generate local transmissions. We extended the analysis to all imported cases combining cases with full and partial information on the timeline. We used the analysis of variance and multiple imputation for the missing data. Results were combined using Rubin’s approach^17^.

### Modeling and predicting importations

We modeled the total number of imported cases out of China over time accounting for date of travel, delay in reporting, and source areas.

We distinguished between three different sources: Hubei province (H), the rest of China (C), other countries (O). We modeled imported cases over time as a piecewise exponential function depending on the source and on travel restrictions in place. We assumed a different situation in Hubei province and the rest of the world due to the level of awareness in the different phases of the outbreak. The exponential functions are defined as follows:

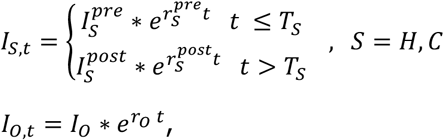

where 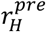 is the growth rate of cases coming from Hubei, and 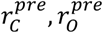, with 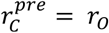, the growth rates of cases coming from the rest of China and other countries, respectively. Travel restrictions were modelled by assuming a discontinuity in the growth rate. For Hubei, we assumed the growth rate to change from 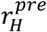 to 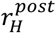 after the travel ban of January 23, 2020 (indicated with *T*_*C*_); for the rest of China, we assumed an analogous change from 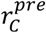 to 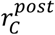 after January 29, 2020 (*T*_*C*_), date of first flight cancellations^18^. No change was considered for the other countries (*r*_0_ constant over time), as no restrictions of travel were established towards these countries. The scale parameters of the exponential functions 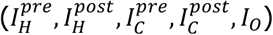 were assumed to be different among the three sources, to account for different traveling volumes and dates of beginning of importations.

We modelled the time *τ* from importation to detection of a case with a gamma distribution, *g*_*t*_(*τ*), conditioned to the date of case importation, *t. g*_*t*_(*τ*) was assumed to have constant coefficient of variation (SD/mean) achieved by a constant shape parameter and a rate parameter varying smoothly in time to capture change in surveillance efficiency.

We used a Bayesian framework to fit the model to imported cases by origin, travel date, and confirmation date. Cases with partial information (e.g. missing date and/or origin of travel) were included by defining latent variables marginalized out during inference. The model was then used to nowcast imported cases two weeks in the future. All details of the analysis are reported in the Appendix.

### Estimation of under-detection of imported cases

We analyzed clusters of transmission generated by imported cases (index case(s) in each cluster) to estimate undetected importations. A cluster can be seeded by more than one index case when local transmissions are epidemiologically linked to more cases traveling together (e.g. infected family members traveling together). We modelled the number of such ‘cluster seeds’, i.e. groups of index cases, with a multinomial distribution depending on the portion of cluster seeds of size 1 or greater than 1 (for simplicity, this was taken as 2), on the probability of detection of a seed, and on occurrence of secondary transmission. The likelihood function was a function of: the number *x*_2_ of observed clusters with one index case; the number *x*_1_ of observed clusters with more than 1 index cases; the number 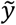 of detected index cases not leading to onward transmission; the number *z* of clusters whose index cases have not been identified; and the number *w* of undetected imported cases that did not generate any cluster. *w* can be estimated through likelihood maximization from the records of 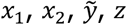.

## RESULTS

### Timeline of travel-related cases

We collected 288 cases, including 163 imported cases, 109 local transmissions, 30 repatriations, and 1 case of unknown origin. Fifteen cases were classified as both imported and local transmissions, since they contracted the infection outside China and traveled to a different country once infected (ES01, ES02, GB03, GB04, GB05, GB06, GB07, GB08, KR12, KR16, KR17, KR19, MY09, TH20, TH21 in our database^16^).

Figure 1 summarizes the timeline of imported cases. Symptoms onset occurred after the travel to the destination country for almost all cases for which date of travel and of onset are available (68 out of 73, 93%). Complete information was available for 51 (31%) imported cases, with quality of information decreasing over time (Figure S1 of the Appendix).

**Figure 1.**
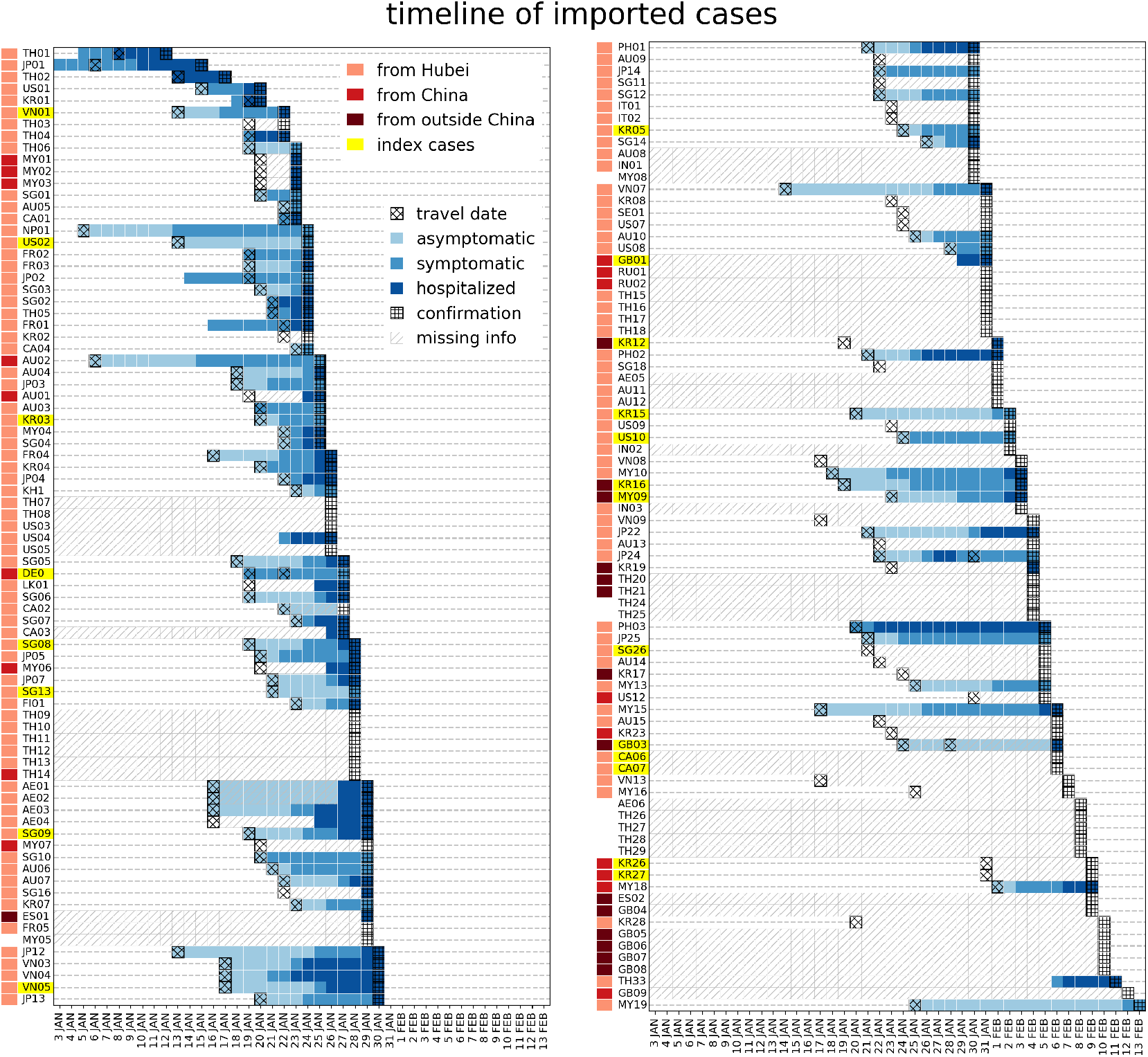
Timeline of importation for all imported cases.

Among imported cases with full information, the delay from travel to hospitalization was longer in cases that generated secondary transmissions (mean of 10 ± 0.97 days compared to 5.5 ± 0.67 days, p=0.003). Overall, the duration from travel to first event (whether symptom onset, or hospitalization for asymptomatic) was also longer, although the difference was not statistically significant (5.0 ± 0.9 days vs. 3.7 ± 0.5 days p= 0.08). Durations of hospitalization were instead comparable among the two groups of cases (1.5 ± 0.7 days vs. 2.6 ± 0.4 days for cases that generated or did not generate secondary transmissions, respectively). Including imported cases with missing information through imputation, we found the same trend though smaller in magnitude and not statistically significant (delay from travel to hospitalization 9.8 ± 1.2 vs. 8.3 ± 0.5 days p= 0.3; delay from travel to onset 5.8 ±1.1 vs. 4.2 ±0.5 p= 0.16, for cases that generated or did not generate secondary transmissions, respectively). This suggests that importations with missing information may be closer in characteristics to index cases leading to onward transmission.

The statistical model predicted a decrease in the average time from travel to detection from 14.5 ± 5.5 days on January 5, 2020 to 6 ± 3.5 days on February 1, 2020 (Figure 2).

**Figure 2.**
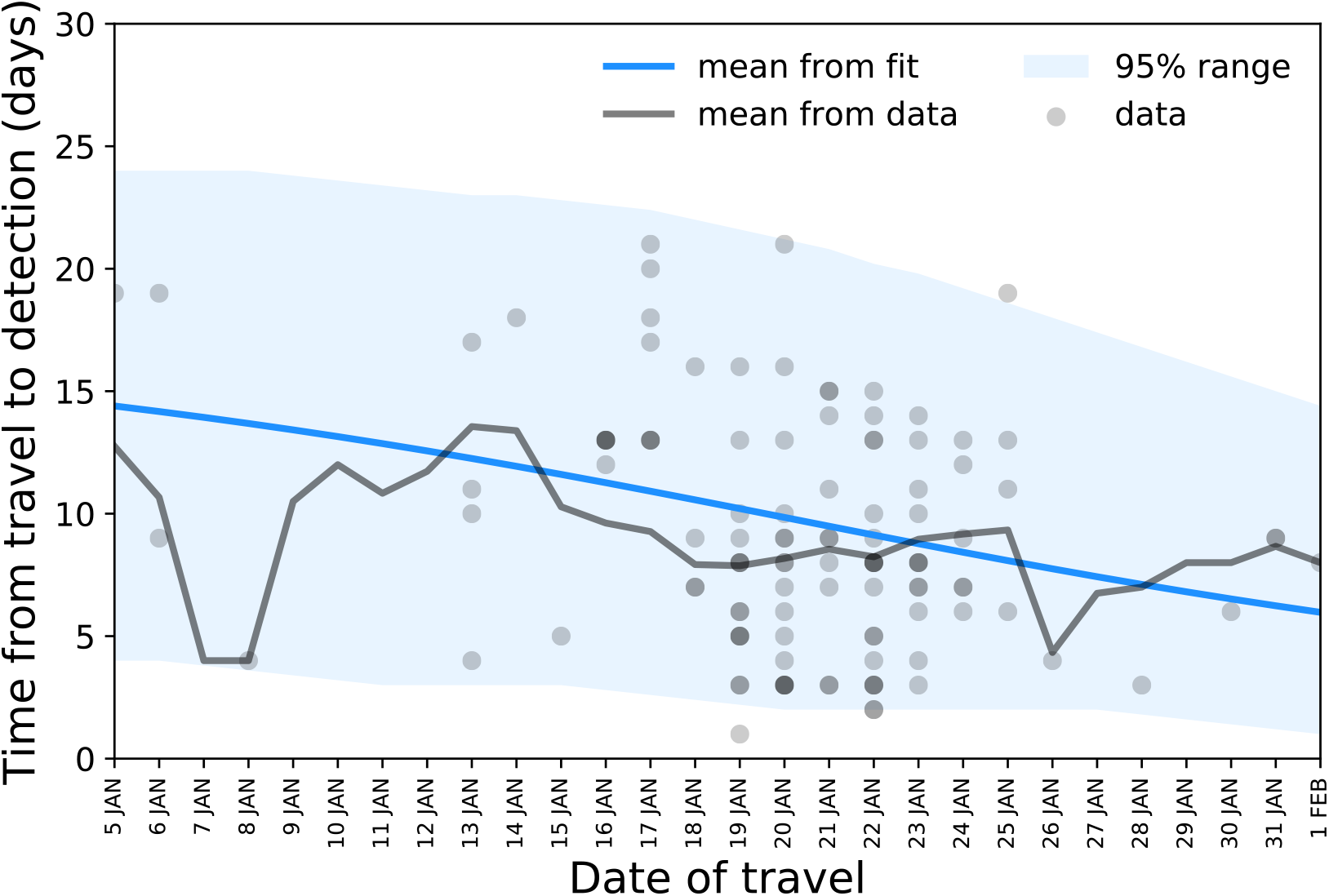
Delay from travel to detection as a function of the date of travel: data points, mean, and model prediction.

### Nowcasting travel-related cases

The model predicted a rapid exponential growth of importations from Hubei, with a growth rate 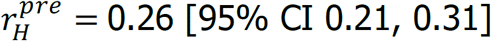, corresponding to a doubling time of 2.8 days. In comparison, the exponential growth from other territories (rest of China and countries other than China) was slow, 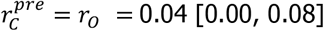. After the implementation of travel restrictions, a negative growth rate was estimated, signaling a decline in imported cases. The decline was sharp for Hubei 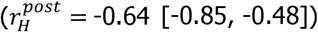 and more gradual for the rest of China 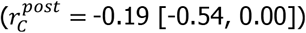.

The predicted trend of all imported cases over time is shown in Figure 3, compared with the observed data. Reported importations are predicted to remain stationary in the second and third week of February and to rise again due to the effect of transmission clusters outside China. Imported cases after February 13, 2020 are in agreement with model predictions (Fig.3).

**Figure 3.**
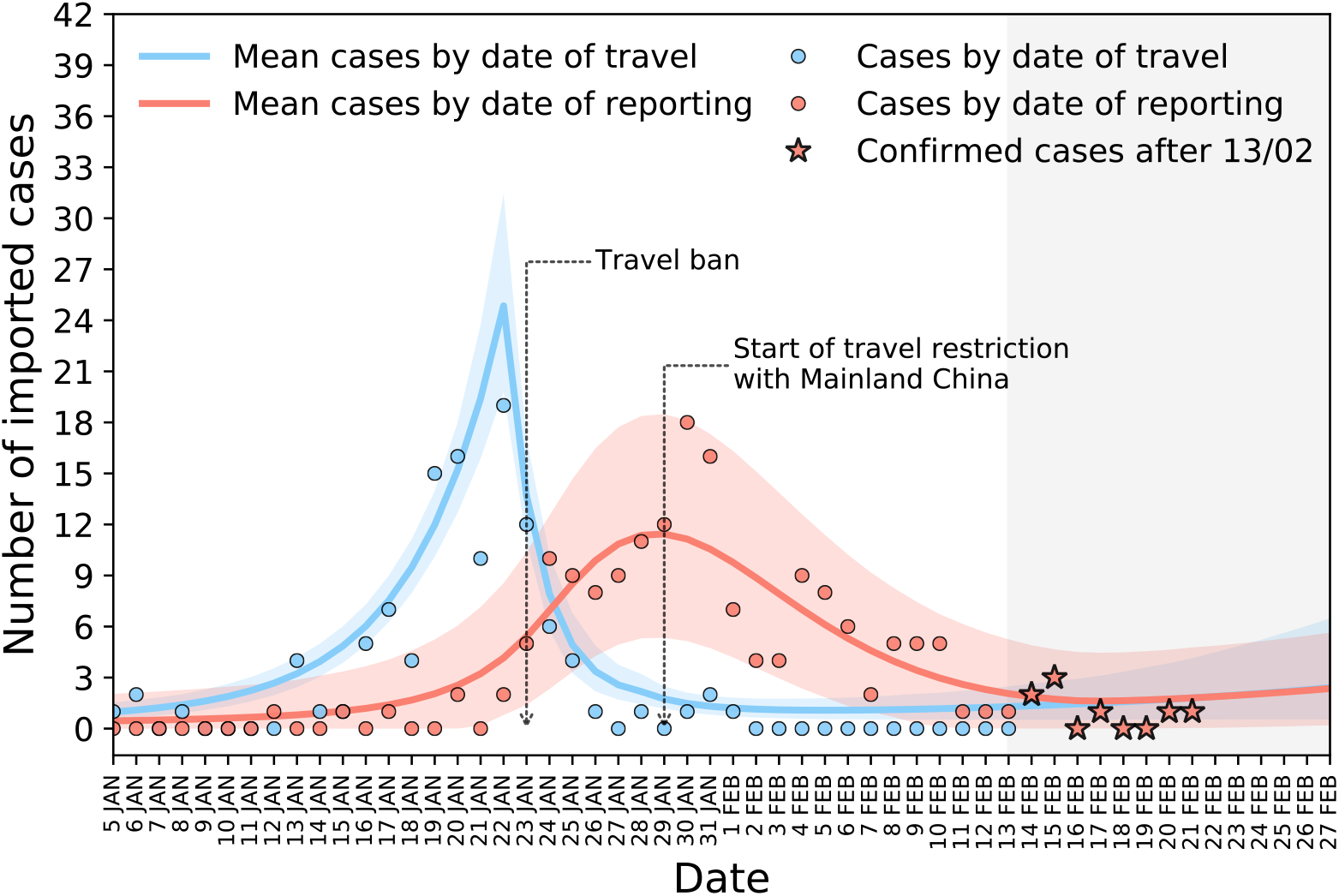
Number of imported cases by date of travel and of reporting: data points and model predictions.

### Trasmission clusters outside China

Forty-two transmission clusters were identified out of China in the timeframe under study. Table 1 summarizes the size and country of each cluster. Clusters were grouped according to whether the index case: (i) was a traveling case identified prior to cluster detection; (ii) a traveling case not identified or identified retrospectively once the cluster was observed; (iii) completely unknown. Assuming that clusters of unknown origin were linked to one of the already observed imported cases - or, in other words, not linked to an undetected imported case - led to an estimate of 76 [49, 118] undetected imported cases. In this scenario, detected cases would amount to 65% of all imported cases. Assuming instead that all clusters of unknown origin were due to undetected imported cases increased the number of undetected cases to 225 [186, 369], i.e. detected cases would correspond to only 36% of the total.

**Table 1.**
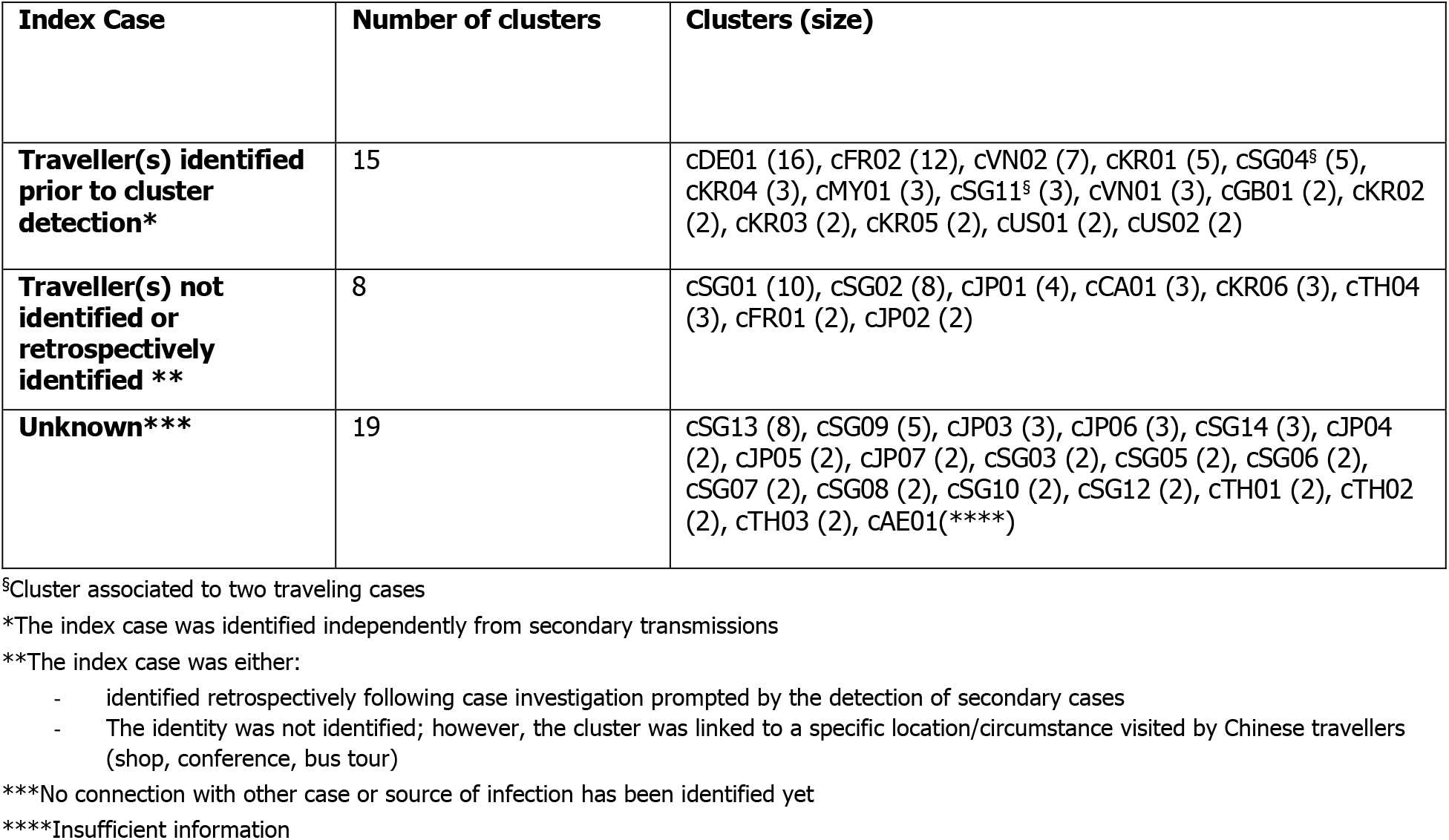
Summary of transmission clusters according to the type of index case.

| Index Case | Number of clusters | Clusters (size) |
| --- | --- | --- |
| **Traveller(s) identified prior to cluster detection\*** | 15 | cDE01 (16), cFR02 (12), cVN02 (7), cKR01 (5), cSG04<sup>§</sup> (5), cKR04 (3), cMY01 (3), cSG11<sup>§</sup> (3), cVN01 (3), cGB01 (2), cKR02 (2), cKR03 (2), cKR05 (2), cUS01 (2), cUS02 (2) |
| **Traveller(s) not identified or retrospectively identified \*\*** | 8 | cSG01 (10), cSG02 (8), cJP01 (4), cCA01 (3), cKR06 (3), cTH04 (3), cFR01 (2), cJP02 (2) |
| **Unknown\*\*\*** | 19 | cSG13 (8), cSG09 (5), cJP03 (3), cJP06 (3), cSG14 (3), cJP04 (2), cJP05 (2), cJP07 (2), cSG03 (2), cSG05 (2), cSG06 (2), cSG07 (2), cSG08 (2), cSG10 (2), cSG12 (2), cTH01 (2), cTH02 (2), cTH03 (2), cAE01(****) |

<sup>§</sup>Cluster associated to two traveling cases
\*The index case was identified independently from secondary transmissions
\*\*The index case was either:
- identified retrospectively following case investigation prompted by the detection of secondary cases - The identity was not identified; however, the cluster was linked to a specific location/circumstance visited by Chinese travellers (shop, conference, bus tour)
\*\*\*No connection with other case or source of infection has been identified yet
\*\*\*\*Insufficient information

## DISCUSSION

As the COVID-19 epidemic in China shows effects of mitigation^2^, increasingly larger clusters of infections reported outside China are raising concern that other territories may start sustaining the outbreak^4,5^. To contain it globally, identification, rapid management of cases, and contact tracing are key. The success of these response measures depends critically on the volume of importations^19^ and the sensitivity of active surveillance^13,15^.

We reviewed here all confirmed cases out of China from January 3 to February 13, 2020 and gathered detailed information on case history and epidemiological links. We identified salient epidemiological features, and modeled the number of importations over time. International exportations from Hubei grew rapidly, fueled by the local epidemic, up to the closure of Wuhan airport preventing further travel of cases. Exportations from other Chinese provinces and other countries grew at a considerably slower pace. This is related to the difference in the increase of cases between Hubei province, origin of the outbreak, and the rest of the affected areas^1^. Such difference is likely an outcome of the implementation of containment measures in China^3,20,21^, and of the increased awareness at different phases of the outbreak^22–26^ (i.e. before and after containment measures) leading to self-isolation and quarantine.

The reduced volume of exported cases worldwide following the travel ban may have given countries the time to prepare and strengthen their surveillance systems, as signaled by a reduction of the interval from travel date to detection over time.

Our model predicts that exportations will likely rise from areas outside China. The number of local transmissions is rapidly increasing in the Republic of Korea, Japan, and Singapore^27^, and few importations in Asia and Europe were registered already from travelers from Japan and Singapore. For this reason, certain countries have updated the history of travel for the case definition of a suspect imported case to include additional countries in South Asia besides China^28^ or banned travelers from East Asian countries^29^. ECDC and WHO currently base their case definition on travel from China only^30,31^, but this may rapidly change in the next days.

Before the likely rebound of exportations, identification and isolation of possible clusters outside China remain essential to contain local transmission. The increasing reporting of clusters outside China with no known epidemiological link^1,14^ raises important concerns on the possibility to contain COVID-19 epidemic worldwide. Our estimates indicate an ability of 36% to detect imported cases in countries outside China. This means that approximately 6 imported cases out of 10 have gone undetected. Previous estimates range from 27%^13^ to 38%^13,15^ detection rates, with variations across countries^13,15^. Ascertainment was estimated to be even lower (approximately 10%) when assessed on repatriations^31^. Here, we excluded from this analysis all repatriation events and cruises with outbreaks, as conditions for detection and identification may be different.

Underdetection may be due to several different factors including asymptomatic infections, infections with mild clinical symptoms, health-seeking behavior and declaration of travel history, case definition, and underdiagnosis. Underdetection of imported cases is likely to be higher than what we estimate here, as our analysis is conditional to the identification of clusters of cases. The current situation in Italy, with different clusters emerging in the timeframe of few hours in different areas in the North of the country^14^, shows that clusters have gone undetected and epidemiological links with the index case are still missing. Countries outside China should be prepared for the possible emergence of several undetected clusters of chains of local transmissions. Surveillance efforts to track all suspect cases may become impractical if the number of cases increases too rapidly^32^. If that situation occurs, countries should be ready to step-up their response and take preparatory steps for community interventions.

## Data Availability

Data were collected by the authors and made publicly available online.

https://docs.google.com/spreadsheets/d/1X_8KaA7l5B_JPpwwV3js1L6lgCRa3FoH-gMrTy2k4Gw/edit#gid=486974430

## ACKNOWLEDGMENTS

This study is partially funded by: the ANR project DATAREDUX (ANR-19-CE46-0008-03); the EU grant MOOD (H2020-874850); the Municipality of Paris through the programme Emergence(s). We thank REACTing (https://reacting.inserm.fr/) for useful discussions.

## APPENDIX

### 1. DATA

**Table S1.** Official sources for international cases

|  |  |
| --- | --- |
| WHO | https://www.who.int/emergencies/diseases/novel-coronavirus-2019/situation-reports/ |
| ECDC | https://www.ecdc.europa.eu/en/geographical-distribution-2019-ncov-cases |
| Victoria Department of Health | https://www2.health.vic.gov.au/about/media-centre/mediareleases# |
| Queensland Public Health | https://www.health.qld.gov.au/news-events/health-alerts/novel-coronavirus |
| Toronto Public Health | https://www.toronto.ca/community-people/health-wellness-care/diseases-medications-vaccines/coronavirus/ |
| Emirates News Agency | https://www.wam.ae/en |
| Bavarian State Ministry of Health | https://www.stmgp.bayern.de/ministerium/presse/ |
| French Ministry of Health | https://solidarites-sante.gouv.fr/ |
| UK Government Public Health | [https://www.gov.uk/health-and-social-care/public-health#news and communications](https://www.gov.uk/health-and-social-care/public-health#news-and-communications) |
| Italian Ministry of Health | http://www.salute.gov.it/portale/nuovocoronavirus/homeNuovoCoronavirus.html |
| India Ministry of Health | https://mohfw.gov.in/ |
| Japan Ministry of Health | https://www.mhlw.go.jp/index.html |
| KCDC Press Release | https://www.cdc.go.kr/board/board.es?mid=a30402000000&bid=0030 |
| Malaysia Ministry of Health | http://www.moh.gov.my/index.php/pages/view/349?mid=29 |
| Philippines Department of Health | https://www.doh.gov.ph/ |
| Russian Government | http://government.ru/en/news/ |
| Public Health Agency of Sweden | https://www.folkhalsomyndigheten.se/the-public-health-agency-of-sweden/communicable-disease-control/novel-coronavirus-2019-ncov/ |
| CDC | https://www.cdc.gov/media/dpk/diseases-and-conditions/coronavirus/coronavirus-2020.html |
| Singapore Ministry of Health | https://www.moh.gov.sg/2019-ncov-wuhan |
| Thailand Ministry of Public Health | https://pr.moph.go.th/?url=pr/index/2/04 |
| VnExpress Health News | https://vnexpress.net/suc-khoe |

### 2. STATISTICAL METHODS

#### Modelling traveling cases and delay from arrival to detection

##### Dataset

The individual data consists of t-uples (*S, f, o*), where:

- *S* indicates place of departure as Hubei province (*H*), China other than Hubei (*C*), outside China (*O*);
- *f* ∈ {1, …, *T*} is the day the case arrived at destination, counted from January 5^th^ up to current date *T*;
- *o* ∈ {1, …, *T*} is the day the case was confirmed, counted from January 5^th^.

##### Modelling the detection delay

The difference *D* = *o* − *f* corresponds to the time from arrival to confirmation. To account for changes in detection efficiency, we modelled *D* as a (discretized) gamma distribution with parameters changing with time. More precisely, the rate parameter of the distribution was *β*_*f*_ = *a* ∗ *e*^*bf*^. The shape parameter *k* was constant, leading to a constant coefficient of variation (Standard deviation/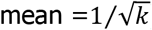).

We truncated the distribution at *T*_*D*_ = 25 days and computed probabilities that *D* was τ days as:

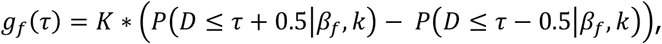

where *K* is a normalization constant accounting for the truncation at *T*_*D*_.

We denote the corresponding cumulative distribution function of D by *G*_*f*_(*τ*) = *P*(*D* ≤ *τ* + 0.5|*β*_*f*_, *k*).

##### Modelling cases arrival

We computed *A*_*S*_ = {*A*_*S,t*_}_*t*=1,…,T_ the daily number of cases arriving from location *S* on date t that had been detected before time T, and *N*_*S*_ = ∑_*t*_ *A*_*S,t*_ the total number of such cases arriving from location *S*.

Due to the time lag between arrival and confirmation, some cases arriving on time *t* from location *S* can be undetected as of time T. We denote *U*_*S,t*_ the number of such cases. Then, the total count of cases arriving on day *t* is given by *A*_*S,t*_ + *U*_*S,t*_. We assumed a Poisson distribution for this count, *A*_*S,t*_ + *U*_*S,t*_∼ *Poisson*(*I*_*S,t*_), where *I*_*S,t*_ represents the expected number of imported cases from location *S* on day *t*.

We modelled *I*_*S,t*_ as a piecewise exponential function in each location of origin S, the exponential growth parameter changing in Hubei after the ban instated on January 23^rd^ and in the rest of China after flight cancellation by major airline companies on January 29^th^. *I*_*S,t*_ was therefore:

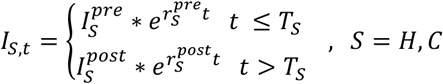

where *T*_*S*_ is the last day *before* the start of quarantine/travel restriction in location 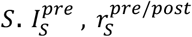 are hyperparameters representing the scale and the growth rate of each exponential, and 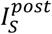 is determined by continuity of *I*_*S,t*_ at *T*_*S*_.

Outside China we assumed a single exponential function with the same growth rate as in China outside Hubei before travel restrictions were put in place 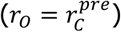 and a different scale :

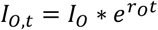

49 confirmed cases had no information on date of arrival and/or origin of travel. These cases were described with latent variables as follows:

- 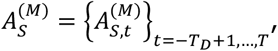, the time series that accounts for case counts with unknown date of arrival;
- 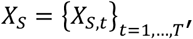 i.e. case counts with unknown travel origin;
- 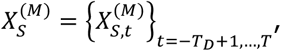, i.e. cases with both information missing.

The framework described above was extended to account for these cases, i.e. we considered 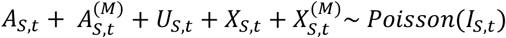 to be the number of cases arriving from destination *S* on time

##### Likelihood function

The components of the estimated parameters θ and prior distributions are listed in Table S2.

**Table S2.** summary of parameters and their priors.

| Parameter | Description | Prior distribution |
| --- | --- | --- |
| $I_H^{pre}$ | Scaling factor Hubei | $\log I_H^{pre} \sim \text{Normal}(0,1)$ |
| $I_C^{pre}$ | Scaling factor China | $\log I_C^{pre} \sim \text{Normal}(0,1)$ |
| $I_o$ | Scaling factor outside China | $\log I_o \sim \text{Normal}(0,1)$ |
| $r_H^{pre}$ | Pre-ban growth rate from Hubei | $\text{Normal}(0,1)$ |
| $r_H^{post}$ | Post-ban growth rate from Hubei | $\text{Normal}(0,1)$ |
| $r_C^{pre}$ | Pre-ban growth rate from China (no Hubei) | $\text{Normal}(0,1)$ |
| $r_C^{post}$ | Post-ban growth rate from China (no Hubei) | $\text{Normal}(0,1)$ |
| $k$ | Shape parameter in time from arrival to detection distribution | $\chi^2(4)$ |
| $a$ | Scale hyperparameter in time from arrival to detection distribution | $\text{Exponential}(1)$ |
| $b$ | Scale hyperparameter in time from arrival to detection distribution | $\log b \sim \text{Normal}(0,1)$ |

The likelihood of the observations is given by:

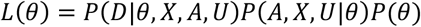

Where:

- *P*(*A, X, U*|*θ*) is the term describing observed incidence according to the model as:

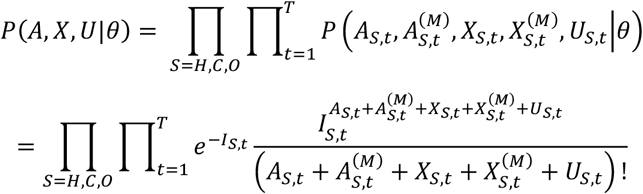

where *I*_*S,t*_ is the expected incidence in location *S* at day *t* described above;
- *P*(*D*|*θ*) is the term describing observed and unobserved duration between arrival and detection:

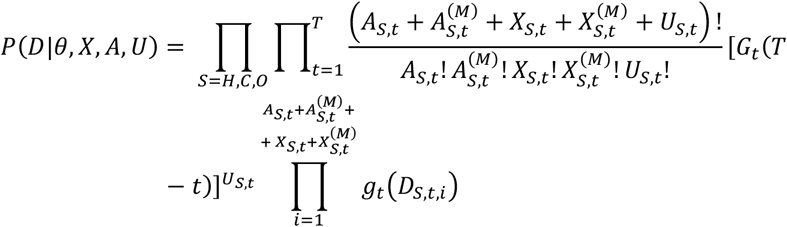

where *D*_*S,t,i*_ are the individual times to detection of those travelling from location *S* on day *t*,

*-P*(*θ*) is the prior model for all parameters

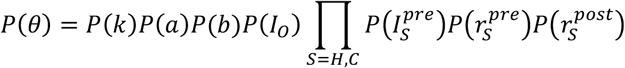

For ease of computation, the likelihood is marginalized over latent variables 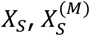 and 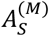 corresponding to cases with missing information 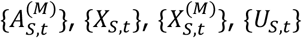, so that data augmentation is unnecessary in the computation of the posterior distribution for the parameters. The final likelihood is:

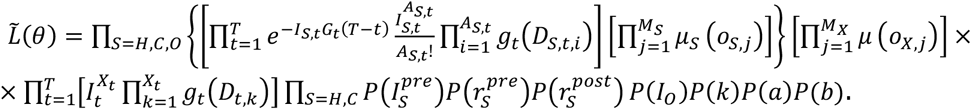

Here we have defined for convenience the following variables: 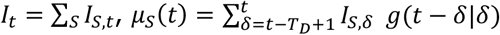 and *μ*(*t*) = Σ_*S*_ *μ*_*S*_(*t*) and introduced *M*_*S*_ the number of cases travelling from source *S* and with unknown date of arrival, *X*_*t*_ is the number of cases that arrived on day *t* from an unknown source, and *M*_X_ is the number of cases with unknown travel source and date of arrival.

Inference was performed by MCMC sampling using *Stan*. We used 3 chains with 6000 iterations and discarded the first 50% as burn-in.

We computed the median of the posterior distributions as well as credible intervals for each parameter in θ. Additionally, we computed predictive distribution statistics about the number of cases confirmed on day *t*, e.g. the average value as well as upper and lower quantiles, using Poisson distribution with mean *μ*(*t*) = ∑_*S*= *H,C,O*_*μ*_*S*_(*t*).

#### Modelling index case detection probability

##### Dataset

We define as seed an imported case or a group of cases that could have started a cluster of local transmission outside China. We computed the number *x*_1_ of transmission clusters where a seed of size 1 was among the cases identified in the cluster and likewise *x*_2_ with seeds of size >1. We also computed the number 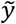 of imported cases that did not start a transmission cluster and the number *z* of clusters for which a seed was not observed among the tested cases, i.e. clusters without a direct link to an imported case.

##### Modeling index case detection

We assumed that seeds could be of size 1 with probability *λ* or of size 2 with probability 1 − *λ*. A seed could be observed with probability *π* and started a cluster with probability *φ*.

The number 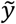 of imported cases that did not start a cluster consist of *y*_*1*_ and *y*_*2*_ seeds of size 1 and 2 such that 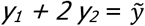 and *y*_*1*_ *+ y*_*2*_ *= y*; however the grouping of these cases is unknown. We computed *y* out of 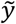 using a plug-in estimate where the mean of the fraction *y*_*1*_*/y*_*2*_ was *λ*/(1 − *λ*), i.e 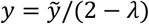.

Denote *w* the number of seeds of any size that went undetected and did not give start to a cluster, with probability: (1 − *π*)(1 − *φ*). *w* is latent and estimated together with *λ, π* and *φ*.

##### Likelihood function

The likelihood is based on a multinomial distribution for *x*_1_, *x*_2_, *y, z* and *w*:

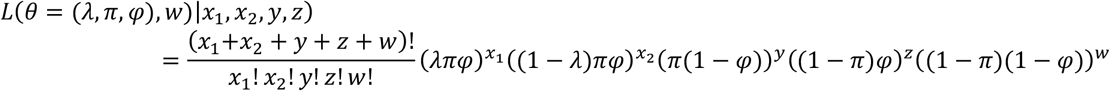

Parameters can be estimated at maximum likelihood:

- Differentiating the likelihood function according to *λ, π* and *φ*:

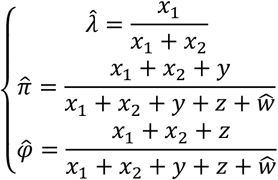
- Approximating the maximum *w* by looking for the value where *L*(*θ, w*) = *L*(*θ, w* − 1) (Pollock KH, Building Models of Capture-Recapture Experiments, The Statistician (1976); 25 (4) : 253-9). We then find:

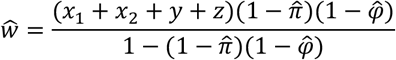

By replacing 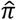 and 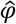 in the previous equation we find that the Maximum Likelihood estimator for *w* is given by:

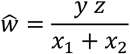

Confidence intervals are computed using profile likelihood methods.

Finally, we estimate the number of unobserved cases that did not give start to a cluster as 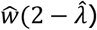. The confidence interval on this last quantity is computed by multiplying the confidence intervals of both factors.

### 3. ADDITIONAL RESULTS

#### Dataset of international cases

We analyze in Figure S1 the proportion of traveling cases for which we have complete information regarding the timeline of events. Detailed information of the clusters of transmission is reported in Table S3.

**Figure S1.**
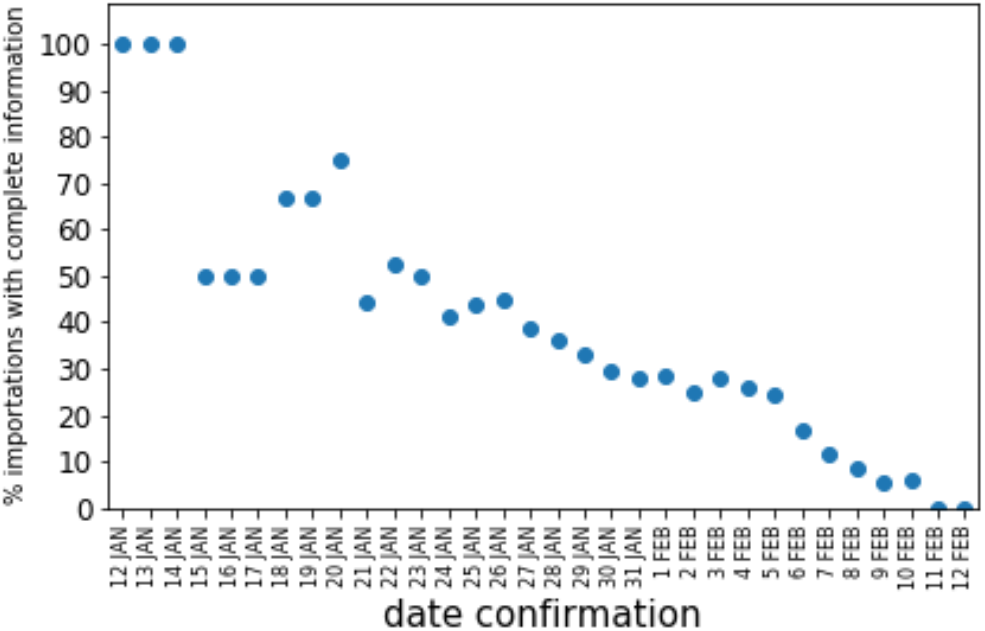
Fraction of imported cases for which we have a complete information on the timeline of importation.

**Table S3.**
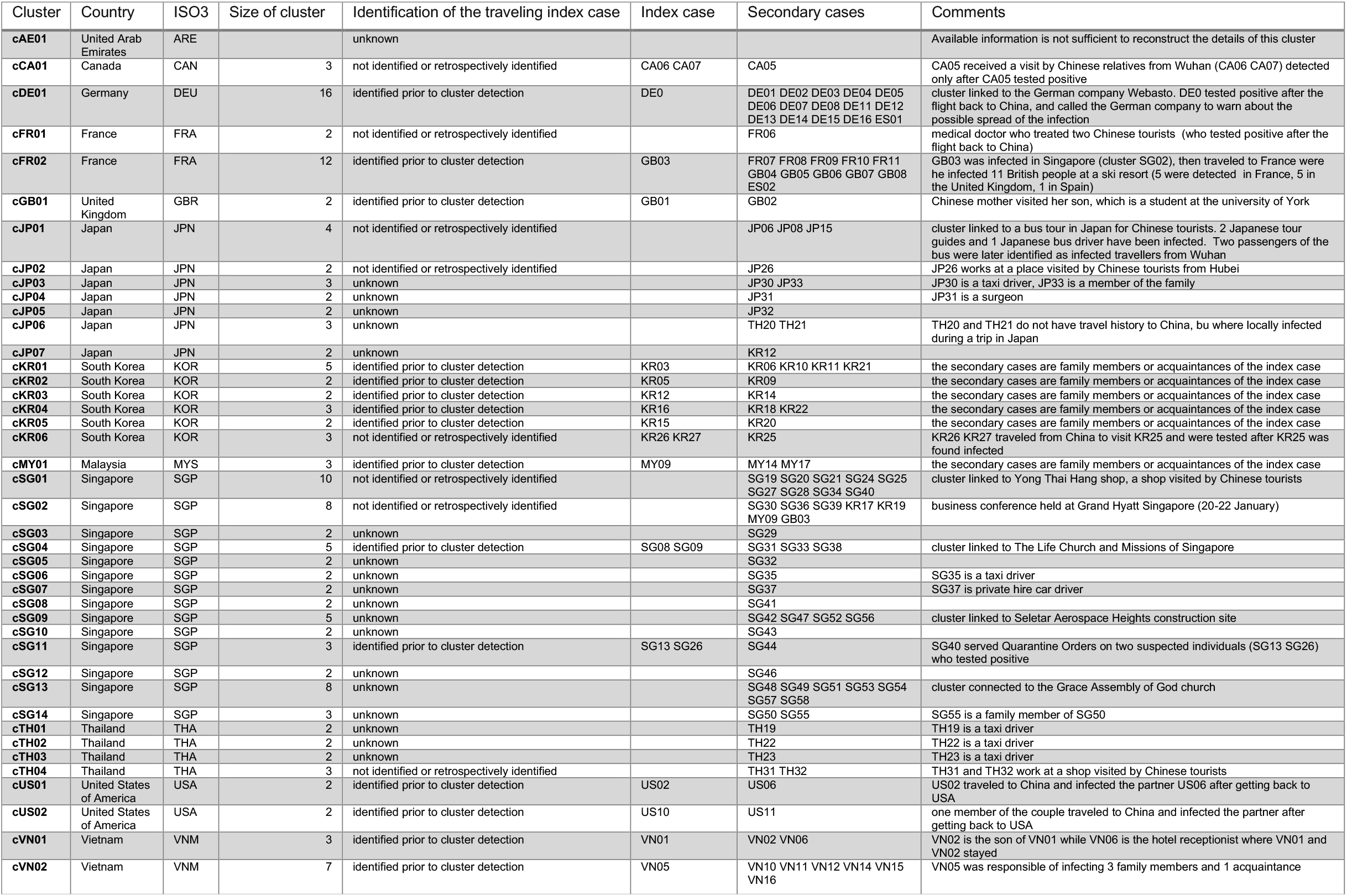
Clusters of local transmission

#### Results of likelihood estimation

We provide in Table S4 all parameter estimates and their confidence intervals. The convergence of the MCMC and the posterior distribution of key parameters are shown in Figure S2.

**Table S4.** Summary of parameter estimates

| Parameter | Median | 95% C. I. |
| --- | --- | --- |
| $I_H^{pre}$ | 0.23 | (0.1, 0.50) |
| $I_C^{pre}$ | 0.38 | (0.19, 0.75) |
| $I_O$ | 0.22 | (0.09, 0.51) |
| $r_H^{pre}$ | 0.26 | (0.21, 0.31) |
| $r_H^{post}$ | -0.64 | (-0.85, -0.48) |
| $r_C^{pre}$ | 0.04 | (0.00, 0.08) |
| $r_C^{post}$ | -0.19 | (-0.54, 0.00) |
| $k$ | 3.32 | (2.55, 4.22) |
| $a$ | 0.16 | (0.08, 0.25) |
| $b$ | 0.05 | (0.02, 0.07) |

**Figure S2.**
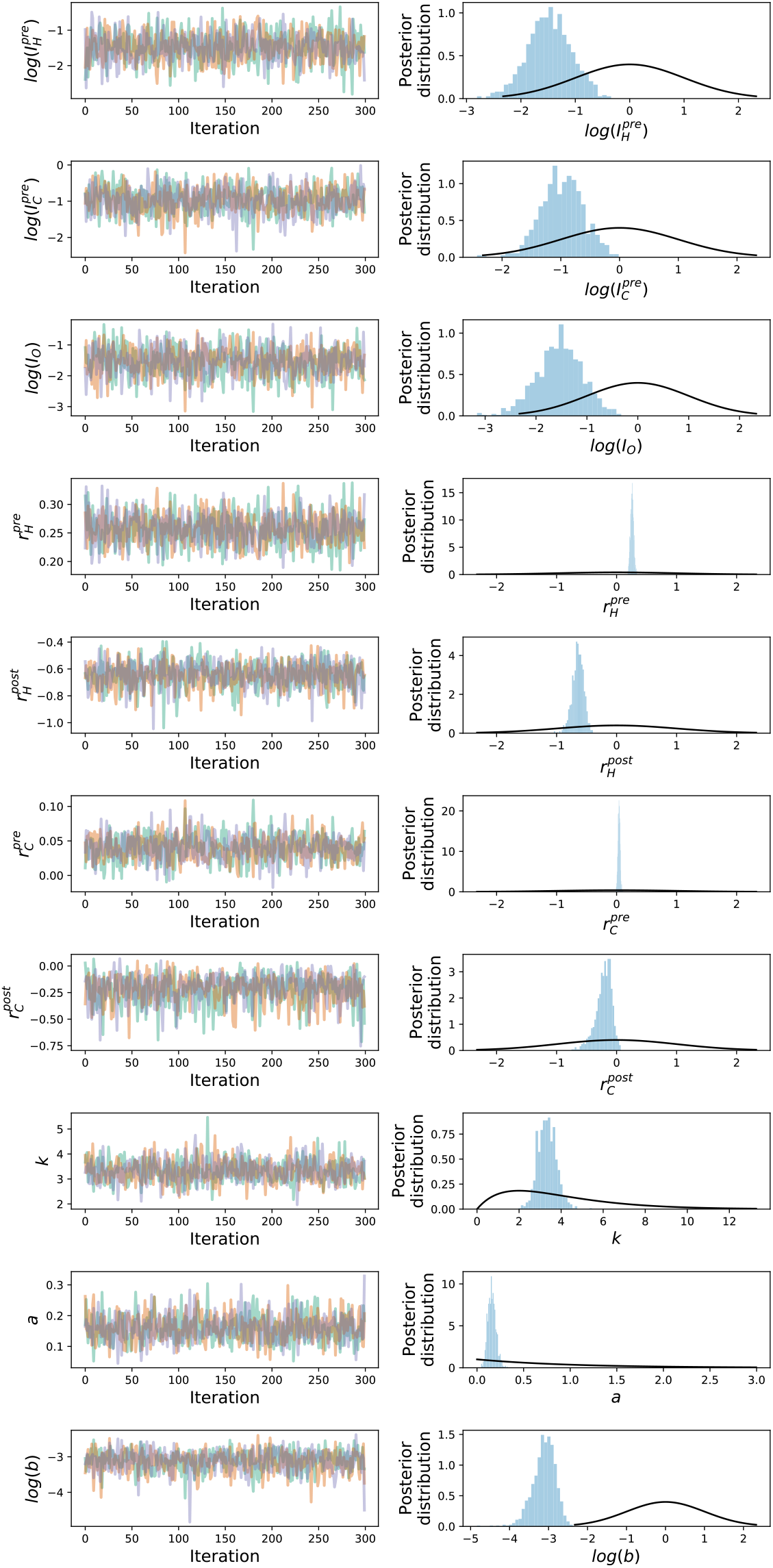
Convergence of MCMC fitting procedure. On the left we show the evolution of each chain for every individual parameter. On the right we plot the corresponding posterior distribution (shaded histogram) as well as the posterior distribution (black line).

#### Sensitivity analysis

On 23/01/2020 all trains, flights and public transports connecting Wuhan with the outside were suspended. We accounted for the possibility that this ban was initially not completely effective, e.g. people at the point of departing were still able to get out of the area with private transports. We consider a sensitivity scenario in which the effects of the travel ban in Wuhan took place on the 24/01/2020, one day later. We found that growth rates changed slightly with respect to the baseline case; in particular 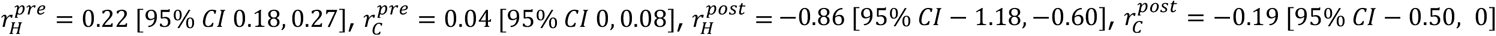.

#### Analysis of imported clusters: summary of parameter estimates

Here we report Maximum Likelihood estimates of parameters in the analysis of imported clusters. We estimate the number of unobserved cases that did not give start to a cluster as 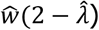. The confidence interval on this last quantity is computed by multiplying the confidence intervals of both factors. For *z* = 8 and *z* = 27 we estimate 76 [49, 118] and 255 [186, 369] undetected cases, respectively. We then estimate the fraction of detected imported cases as 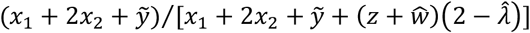, which yields 65% and 36% for *z* = 8 and *z* = 27, respectively.

**Table S5.**
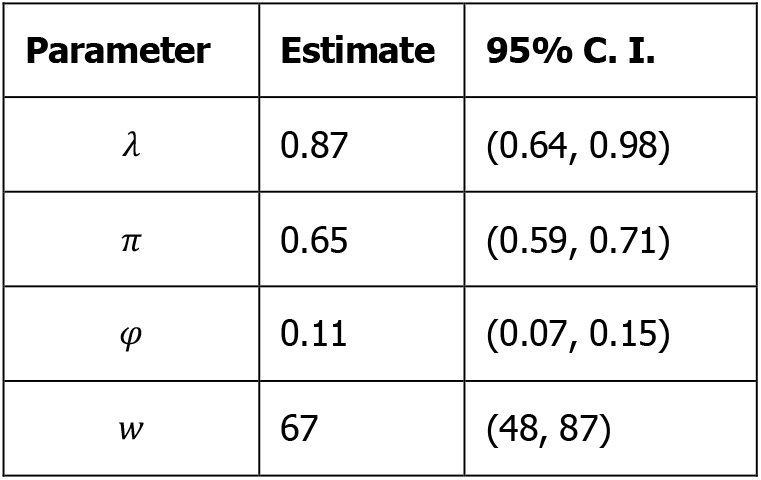
Summary of parameter estimates for *x*_1_ = 13, *x*_2_ = 2, *ỹ* = 142, *z* = 8.

**Table S6.**
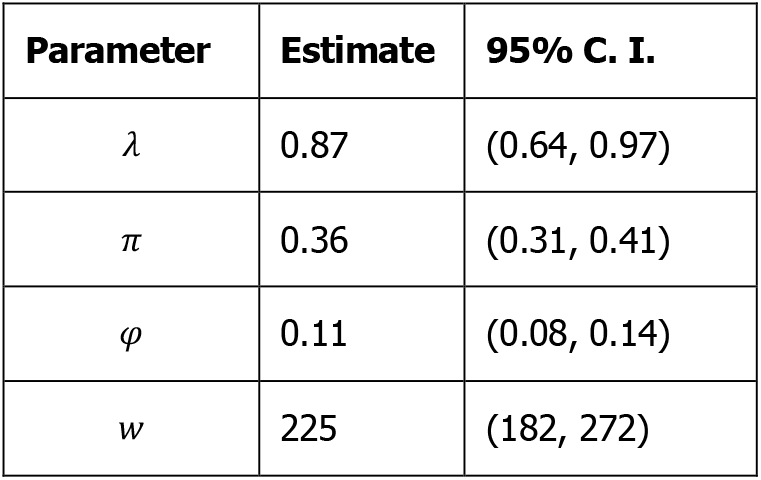
Summary of parameter estimates for *x*_1_ = 13, *x*_2_ = 2, *ỹ* = 142, *z* = 27.

## Notes

### Competing Interest Statement

The authors have declared no competing interest.

### Funding Statement

This study was partially funded by: the ANR project DATAREDUX (ANR-19-CE46-0008-03); the EU grant MOOD (H2020-874850); the Municipality of Paris through the programme Emergence(s).

## REFERENCES

1. World Health Organization (WHO). Coronavirus disease (COVID-2019) situation reports - 31. https://www.who.int/emergencies/diseases/novel-coronavirus-2019/situation-reports.

2. Novel Coronavirus Pneumonia Emergency Response Epidemiology Team. [The epidemiological characteristics of an outbreak of 2019 novel coronavirus diseases (COVID-19) in China]. Zhonghua Liu Xing Bing Xue Za Zhi Zhonghua Liuxingbingxue Zazhi 41, 145–151 (2020).

3. Tian, H. et al. Early evaluation of transmission control measures in response to the 2019 novel coronavirus outbreak in China. medRxiv 2020.01.30.20019844 (2020) doi:10.1101/2020.01.30.20019844.

4. World Health Organization (WHO) on Twitter: ‘@DrTedros @_AfricanUnion @NCDCgov @AfricaCDC @JNkengasong @UN @antonioguterres @DrMikeRyan SThe increasing signs of transmission outside #China show that the window of opportunity we have for containing this #coronavirus is narrowing. We are calling on all countries to invest urgently in preparednessS-@DrTedros #COVID19’ / Twitter. Twitter https://twitter.com/who/status/1231112868608823298.

5. World Health Organization (WHO) on Twitter: ‘@DrTedros @_AfricanUnion SAlthough the total number of #COVID19 cases outside #China remains relatively small, we are concerned about the number of cases with no clear epidemiological link, such as travel history to CN or contact with a confirmed caseS-@DrTedros #coronavirus’ / Twitter. Twitter https://twitter.com/who/status/1231111164492419072.

6. World Health Organisation. Coronavirus disease (COVID-19) technical guidance. https://www.who.int/emergencies/diseases/novel-coronavirus-2019/technical-guidance.

7. Heymann, D. L. & Shindo, N. COVID-19: what is next for public health? The Lancet 395, 542–545 (2020).

8. Gostic, K., Gomez, A. C. R., Mummah, R. O., Kucharski, A. J. & Lloyd-Smith, J. O. Estimated effectiveness of traveller screening to prevent international spread of 2019 novel coronavirus (2019-nCoV). medRxiv 2020.01.28.20019224 (2020) doi:10.1101/2020.01.28.20019224.

9. Fraser, C., Riley, S., Anderson, R. M. & Ferguson, N. M. Factors that make an infectious disease outbreak controllable. Proc. Natl. Acad. Sci. 101, 6146–6151 (2004).

10. Rothe, C. et al. Transmission of 2019-nCoV Infection from an Asymptomatic Contact in Germany. N. Engl. J. Med. 0, null (2020).

11. Dorigatti, I. et al. Report 4: Severity of 2019-novel coronavirus (nCoV). 12.

12. Kucharski, A. J. et al. Early dynamics of transmission and control of 2019-nCoV: a mathematical modelling study. medRxiv 2020.01.31.20019901 (2020) doi:10.1101/2020.01.31.20019901.

13. MRC Centre for Global Infectious Disease Analysis. Report 6: Relative sensitivity of international surveillance. https://www.imperial.ac.uk/mrc-global-infectious-disease-analysis/news--wuhan-coronavirus/.

14. Salute, M. della. Covid-19 - Situazione in Italia e nel mondo. http://www.salute.gov.it/portale/nuovocoronavirus/dettaglioContenutiNuovoCoronavirus.jsp?lingua=italiano&id=5338&area=nuovoCoronavirus&menu=vuoto.

15. Niehus, R., Salazar, P. M. D., Taylor, A. & Lipsitch, M. Quantifying bias of COVID-19 prevalence and severity estimates in Wuhan, China that depend on reported cases in international travelers. medRxiv 2020.02.13.20022707 (2020) doi:10.1101/2020.02.13.20022707.

16. COVID-19 international cases as of Feb 13. https://docs.google.com/spreadsheets/d/1X_8KaA7l5B_JPpwwV3js1L6lgCRa3FoH-gMrTy2k4Gw/edit?usp=sharing.

17. Rubin, D. B. Multiple Imputation for Nonresponse in Surveys. (Wiley-Interscience, 2004).

18. Business, S. P. and H. Z., CNN. Airlines around the world are suspending flights to China as the coronavirus spreads. CNN https://www.cnn.com/2020/01/29/business/british-airways-coronavirus/index.html.

19. Hellewell, J. et al. Feasibility of controlling 2019-nCoV outbreaks by isolation of cases and contacts. medRxiv 2020.02.08.20021162 (2020) doi:10.1101/2020.02.08.20021162.

20. Chinazzi, M. et al. The effect of travel restrictions on the spread of the 2019 novel coronavirus (2019-nCoV) outbreak. medRxiv 2020.02.09.20021261 (2020) doi:10.1101/2020.02.09.20021261.

21. Lai, S. et al. Assessing spread risk of Wuhan novel coronavirus within and beyond China, January-April 2020: a travel network–based modelling study. medRxiv 2020.02.04.20020479 (2020) doi:10.1101/2020.02.04.20020479.

22. Sun, K., Chen, J. & Viboud, C. Early epidemiological analysis of the 2019-nCoV outbreak based on a crowdsourced data. medRxiv 2020.01.31.20019935 (2020) doi:10.1101/2020.01.31.20019935.

23. Poletto, C., Boëlle, P.-Y. & Colizza, V. Risk of MERS importation and onward transmission: a systematic review and analysis of cases reported to WHO. BMC Infect. Dis. 16, 448 (2016).

24. Rubin, G. J., Amlôt, R., Page, L. & Wessely, S. Public perceptions, anxiety, and behaviour change in relation to the swine flu outbreak: cross sectional telephone survey. BMJ 339, b2651 (2009).

25. Rizzo, C. et al. Survey on the Likely Behavioural Changes of the General Public in Four European Countries During the 2009/2010 Pandemic. in Modeling the Interplay Between Human Behavior and the Spread of Infectious Diseases (eds. Manfredi, P. & D’Onofrio, A.) 23–41 (Springer, 2013). doi:10.1007/978-1-4614-5474-8_2.

26. Majumder, M. S., Kluberg, S., Santillana, M., Mekaru, S. & Brownstein, J. S. 2014 Ebola Outbreak: Media Events Track Changes in Observed Reproductive Number. PLOS Curr. Outbreaks (2015) doi:10.1371/currents.outbreaks.e6659013c1d7f11bdab6a20705d1e865.

27. World Health Organization (WHO). Coronavirus disease (COVID-2019) situation reports - 32. https://www.who.int/emergencies/diseases/novel-coronavirus-2019/situation-reports.

28. COVID-19: guidance for staff in the transport sector. GOV.UK https://www.gov.uk/government/publications/covid-19-guidance-for-staff-in-the-transport-sector/covid-19-guidance-for-staff-in-the-transport-sector.

29. The Times of Israel. Israel bans foreigners coming from East Asian countries over virus fears. https://www.timesofisrael.com/israel-bans-foreigners-coming-from-east-asian-countries-over-virus-fears/.

30. Case definition and European surveillance for human infection with novel coronavirus (SARS-CoV-2). European Centre for Disease Prevention and Control https://www.ecdc.europa.eu/en/case-definition-and-european-surveillance-human-infection-novel-coronavirus-2019-ncov.

31. World Health Organization (WHO). Global Surveillance for human infection with coronavirus disease (COVID-2019). https://www.who.int/publications-detail/global-surveillance-for-human-infection-with-novel-coronavirus-(2019-ncov).

32. Nishiura, H. et al. The Rate of Underascertainment of Novel Coronavirus (2019-nCoV) Infection: Estimation Using Japanese Passengers Data on Evacuation Flights. J. Clin. Med. 9, 419 (2020).

33. Lipsitch, M., Hayden, F. G., Cowling, B. J. & Leung, G. M. How to maintain surveillance for novel influenza A H1N1 when there are too many cases to count. The Lancet 374, 1209–1211 (2009).

